# Multidimensional motoneuron control using intramuscular microelectrode arrays in tetraplegic spinal cord injury

**DOI:** 10.1101/2025.07.17.25331429

**Authors:** Agnese Grison, Ciara Gibbs, Vishal Rawji, Lara Gouveia Vila, Isabella Szczech, Rejin Varghese, Peter Bryan, Aritra Kundu, Xingchen Yang, Juan Álvaro Gallego, Dario Farina

**Author notes:** These authors contributed equally to this work (Agnese Grison, Ciara Gibbs, Vishal Rawji). Co-senior authors (Juan Álvaro Gallego, Dario Farina).

## Abstract

Loss of hand function after spinal cord injury (SCI) significantly impairs independence and quality of life. Although residual muscle activity recorded on the skin can provide intuitive control signals, it is limited in SCI by low amplitude, and poor signal-to-noise ratio. Here, we show that these key limitations can be overcome by implanting 40-channel microelectrode arrays directly into forearm muscles. Using blind source separation methods, we detected the simultaneous activity of up to 73 spinal motoneurons during attempted movements of two tetraplegic individuals with SCI. Even in the presence of spasticity or complete paralysis, participants could modulate the activity of individual motoneurons to accurately control a computer cursor in one and two dimensions, play a video game, and even regain the ability to grasp using a motoneuron-controlled soft exoskeleton glove. Our findings demonstrate the feasibility of an implantable microelectrode array system that uses residual motoneuron activity for control in SCI, with the potential to improve the quality of life for individuals with paralysis.

## Main

Our hands are central to everyday life, enabling interactions with devices such as smartphones, tablets, and computers for socialising, work, entertainment, and engaging with the wider world. For people with tetraplegic spinal cord injury (SCI), loss of hand function significantly impairs independence and quality of life^1,2^. Restoring upper limb function is therefore of high clinical priority.

One promising method to restore functionality involves harnessing residual neural/muscle signals to control computers and assistive devices^3–8^. Electromyographic signals (EMG), generated when action potentials of spinal motoneurons trigger muscle fibre action potentials, is one such signal. Indeed, despite the lesion, EMG is often detectable even after severe SCI^9^. However, due to a paucity of surviving supraspinal connections to spinal motoneurons^10^, EMG can be very small and sparse^11,12^, especially when measured using surface electrodes^13^. This poses a fundamental limitation to stable control using surface EMG in people with SCI. In addition, surface EMG-based systems are limited by the need to reposition electrodes across sessions^14,15^, and by signal variability due to changes in skin-electrode contact, such as those caused by sweat^16^ or movement^17^.

Invasive approaches, by which an electrode is directly inserted into a muscle, increase the signal-to-noise ratio of the recordings, overcoming some of the inherent limitations of surface EMG^18–21^, as well as those specific to people with SCI^22^. However, whilst conventional intramuscular EMG electrodes (e.g., bipolar, multiple fine wires) record high quality signals in healthy individuals, these signals only reflect the activity along a small portion of a muscle^23^. This small detection volume, combined with the limited number of surviving motoneurons in SCI^9^, makes it inherently difficult to detect motoneuron activity in paralysed individuals, and thus alternative approaches are needed.

Here, we present an approach that invasively records from a large muscle volume using 40-channel intramuscular microelectrode arrays^24^. While each electrode site is highly selective, the detection volume is large due to the optimised spatial distribution of the electrodes along a 2 cm long array^24^. Using extensively validated spike sorting methods to extract the activity of spinal motoneurons from these high-density intramuscular recordings^25,26^, we reliably detected the activity of up to 73 motoneurons simultaneously. Remarkably, two tetraplegic individuals could voluntarily control the activity of their motoneurons, achieving up to two-dimensional control of a computer cursor and a soft exoskeleton glove, despite the lack of observable movement. We thus provide proof of feasibility of an implantable microelectrode array system that uses residual motoneuron activity for control in SCI.

## Identification of many independent motoneurons from a single intramuscular microelectrode array

We first checked whether the activity of individual motoneurons could be detected in muscles that showed minimal to no contraction. Participants attempted flexion and extension movements of individual fingers, the thumb and the wrist during ultrasound recordings of forearm muscles (Fig. 1A). By identifying regions of residual fibre contraction, we were able to strategically insert the microelectrode arrays^24^ (Fig. 1B, C) such that all 40 channels laid within contracting muscles (for participant one, two arrays in extensor muscles; for participant two, one array in a flexor muscle and one in an extensor; example insertion in Video 1), obtaining high-dimensional recordings (Fig. 1D).

**Fig. 1:**
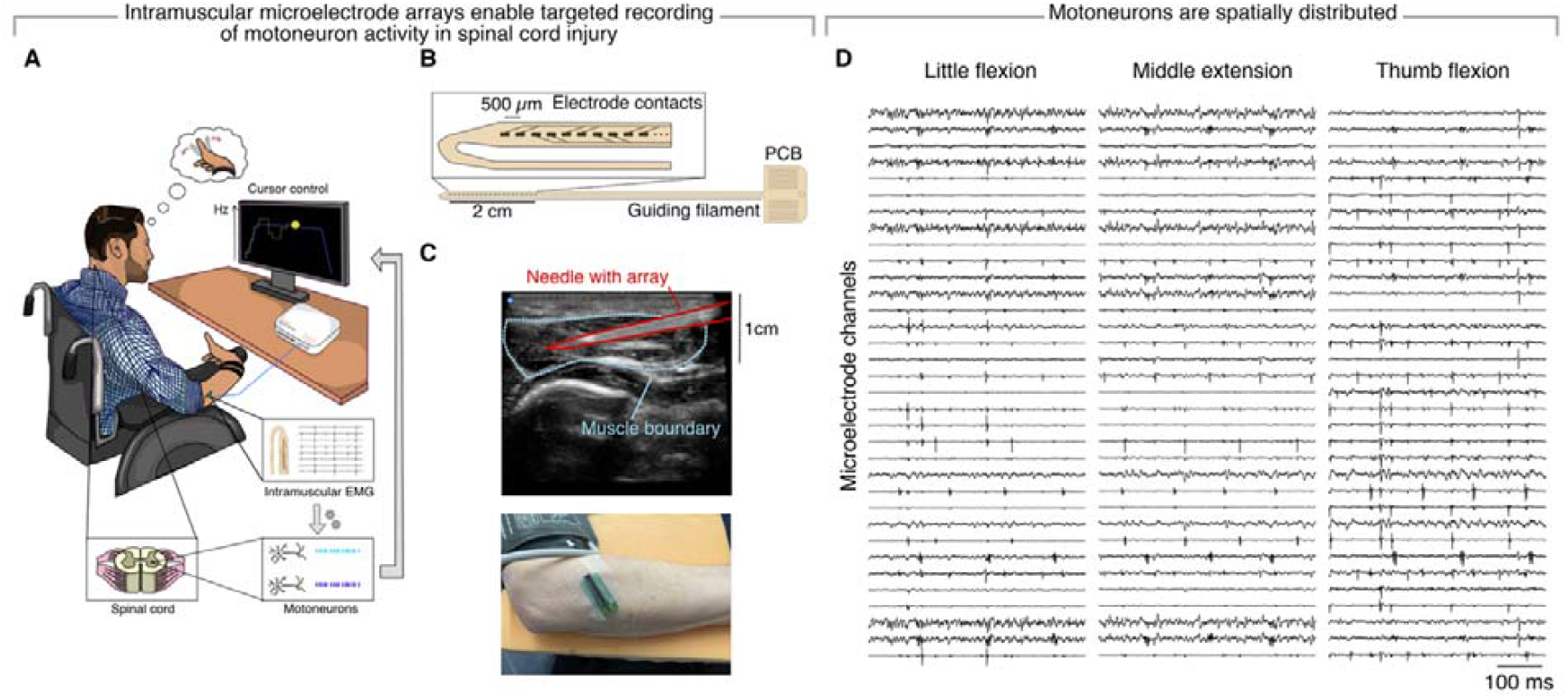
Experimental setup and intramuscular high-density EMG recordings. Overview of the experimental setup, in which participants controlled a computer cursor using the activity of detected motoneurons (**A**). We used high-density microelectrode arrays (**B**), inserted under ultrasound-guidance and connected to a PCB that rested on the surface of the skin (**C**) to record high-dimensional EMG signals. **D** Example recordings during three attempted actions.

Using blind source separation methods^25,26^, we identified the spiking of up to 73 motoneurons for each attempted movement despite the lack of *overt* movement (Fig. 2A,B) (participant one: 11.4 ± 0.8; participant two: 54.2 ± 8.0 per attempted movement; mean firing rate, 8.9 ± 4.9 Hz, and 7.9 ± 2.5 Hz, respectively; values calculated across two microelectrode arrays). Motoneurons were spatially distributed across the microelectrode array according to their action potential waveforms (Fig. 2C,D; Video 2). Motoneuron pairs exhibited low, albeit mostly significant, pairwise correlations in their firing rates (Suppl. Fig. 1; participant one: *r* = 0.12 ± 0.17, 71.2% of the correlations significant; participant two: *r* = 0.10 ± 0.16, 78.7% significant; *p* < 0.05;), which decreased with increasing distance at which the motoneurons were recorded (Suppl. Fig. 1, participant one: 0.19 ± 0.20 vs. 0.02 ± 0.04; participant two: 0.21 ± 0.14 vs. –0.03 ± 0.05). Low correlation values between motoneurons within and between arrays suggest limited shared synaptic inputs, suggesting that they may be independently controllable.

**Fig. 2:**
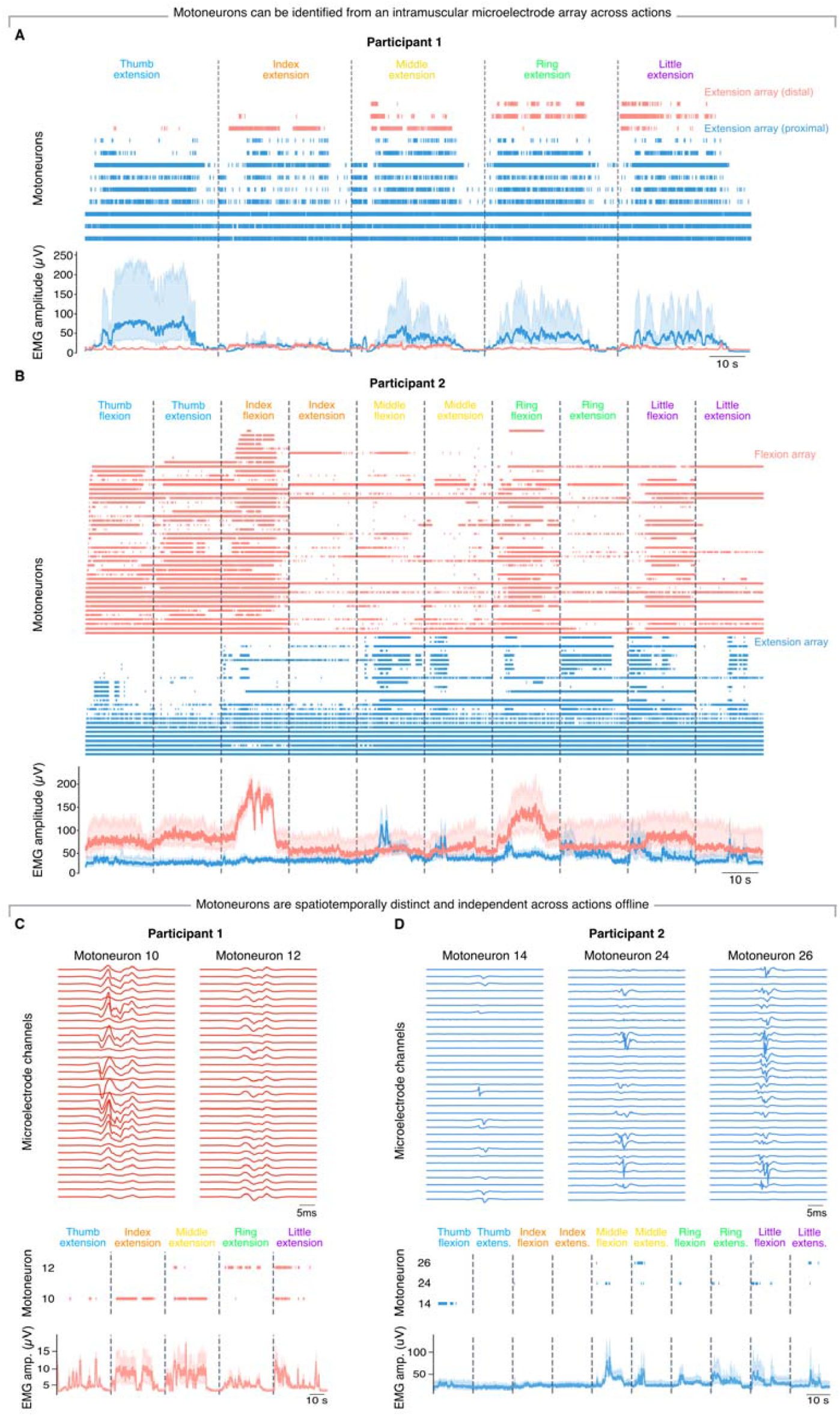
Motoneurons identified during attempted movements. Raster plots show spiking activity of individual motoneurons during attempted movements from participant one (**A**) and two (**B**). Bottom plots, overall EMG amplitude from each array, calculated as the root-mean-squared value. Groups of motoneurons fired independently of one another and were activated by specific attempted movements. **C** Action potentials recorded in the same microelectrode array for two representative motoneurons for participant one. **D** Same for participant two. These independent motoneurons can be distinguished by the spatial distribution of their action potentials across the array.

Whilst most motoneurons only fired during attempted movements (participant one: nine motoneurons, 75.0% of all detected; participant two: 62 motoneurons, 84.9%), others fired continuously during both rest and attempted movements and could not be deactivated (participant one: three motoneurons, 25.0%; participant two: 11 motoneurons, 15.1%). This was evident for both participants, even though only participant two displayed clinical spasticity. Among motoneurons that were under voluntary control, most were active for multiple attempted movements, while a minority were recruited specifically for only one type of attempted movement (participant one: one motoneuron; participant two: eight motoneurons). Both participants had some pairs of motoneurons that were recruited independently of each other when they attempted different movements (Fig. 2C,D) (participant one: two pairs; participant two: 11 pairs), and that could be selectively activated online (Suppl. Fig. 2A,B). We also found a case of three independently activated motoneurons, recruited by three attempted movements, in participant two (Fig. 2D). Remarkably, the same motoneurons could be reliably detected during the entire six hour long experimental session, with those that were controllable remaining under voluntary control for at least four hours (Suppl. Fig. 2C). This was the case even if participants underwent pressure relieving exercises and suffered randomly occurring jerks (myoclonus).

We also recorded high-density surface EMG signals^25^ over the same ultrasound-identified region to compare motoneuron detection to that obtained using intramuscular arrays. Motoneuron yield was far lower with simultaneous high-density surface EMG recordings than with intramuscular microelectrode arrays (compare Suppl. Fig. 3A,B with Fig. 2A,B), despite the larger number of surface EMG channels: only seven motoneurons could be identified non-invasively from participant one, in contrast to the 12 motoneurons identified using intramuscular arrays. In participant two, 18 motoneurons were identified from surface EMG, a yield that was again far fewer than the 73 motoneurons identified using intramuscular arrays. This result highlights the performance of the intramuscular microelectrode array design, which detects the spiking of larger motoneuron numbers thanks to their optimised configuration^18^, outperforming surface EMG techniques even in the case of low amplitude, low signal-to-noise ratio recordings in SCI.

**Fig. 3:**
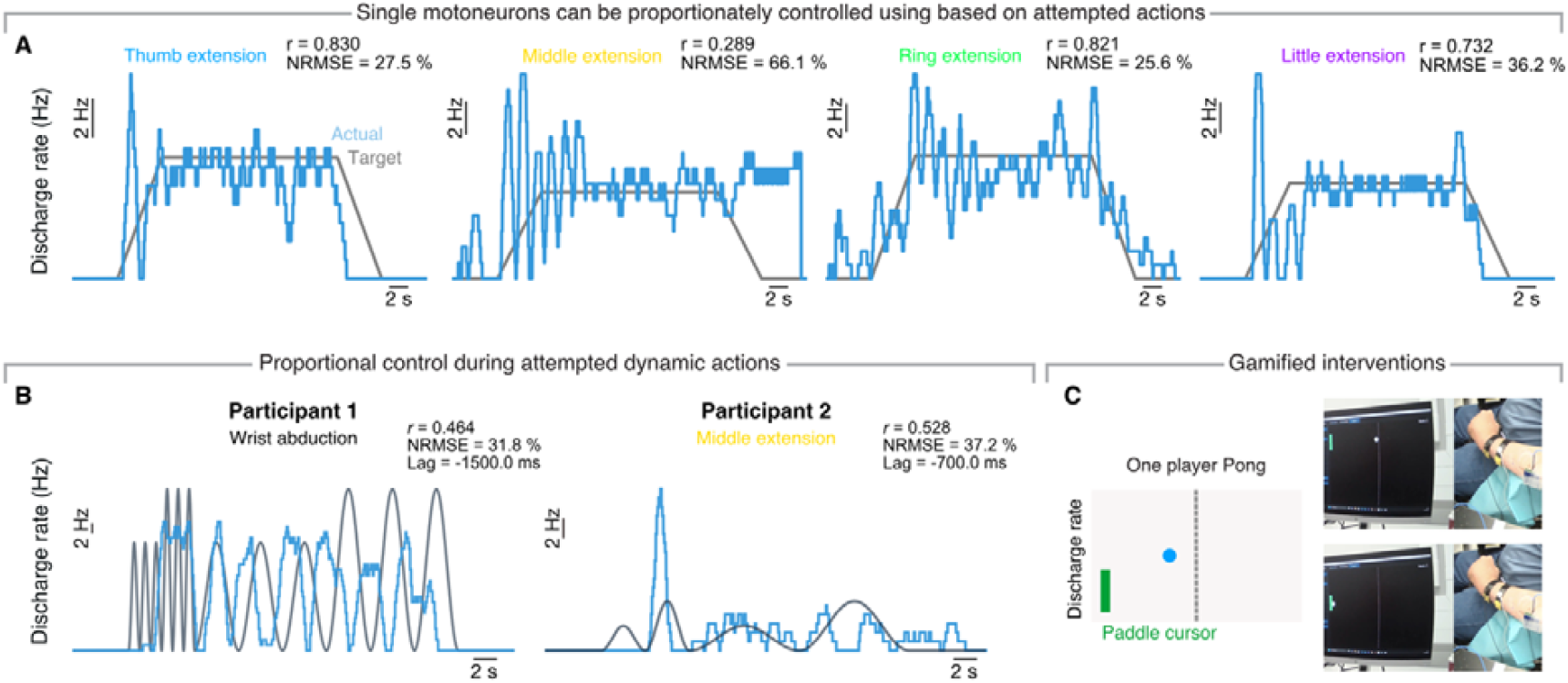
Online one-dimensional motoneuron control. **A** Online tracking of a trapezoidal profile using the firing rate of four different motoneurons each associated with a different attempted action (top). **B** Same for a dynamic profile consisting of sinusoidal components of various amplitudes and frequencies. Time lag represents the optimal temporal offset applied to dynamic task signals to maximise correlation with the target. **C** In Pong, the height of the green paddle increases with motoneuron firing rate. Pictures show control of the paddle to prevent the ball from reaching the left wall.

## Residual motoneuron activity enables accurate, multidimensional control

Proportional modulation of a command signal provides an intuitive way to control an external device or a computer cursor^27–29^. We thus tested whether participants could continuously control the activity of a single motoneuron^30^ to track two distinct profiles, each requiring different control strategies: a trapezoidal profile that required participants to maintain the firing rate of a single motoneuron at a fixed level (Fig. 3A, Suppl. Fig. 4; Video 3), and a dynamic profile that required participants to flexibly modulate the firing rate to different levels (Fig. 3B; Video 4).

**Fig. 4:**
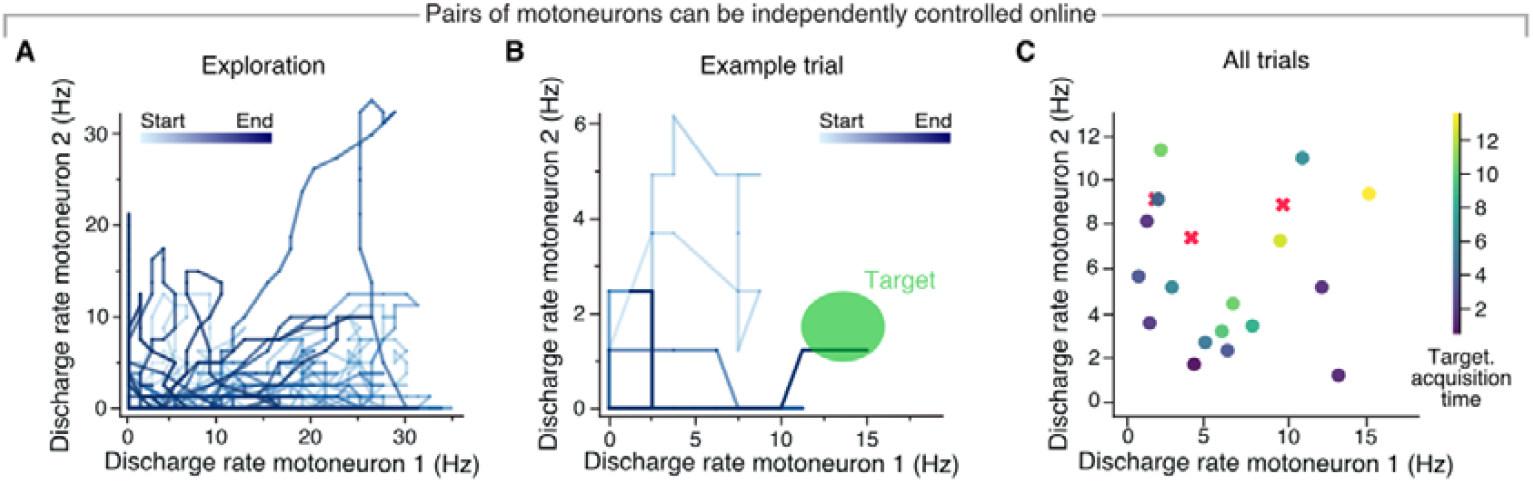
Two-dimensional control using two independent motoneurons. **A**. Example trajectory of a single block of trials (300 s) for participant one, showing that various regions of the workspace could be reached. Colour gradient, time. **B**. Example of a target successfully reached during the experiment. Colour gradient, time. **C**. Targets and acquisition time (colour) for an example session from Participant 1. X indicates failure.

Both participants successfully modulated the firing rate of a single motoneuron to track trapezoidal and dynamic profiles (Fig. 3A,B, Videos 3 and 4). Performance on the trapezoidal profile was varied: while participant two was able to sustain a stable discharge for the entire 20 s plateau (Fig. 3A; trapezoids: *r* = 0.83 ± 0.004, NRMSE = 26.6% ± 0.9%), participant one struggled to maintain the discharge rate at a constant level (Suppl. Fig. 4; *r* = 0.38 ± 0.02, NRMSE = 60.3% ± 3.6%). In contrast, participants performed similarly well during dynamic tracking (Fig. 3B; participant one: *r* = 0.47, NRMSE = 31.8%, optimal time lag = −1.5 s; participant two: *r* = 0.53, NRMSE = 37.2%, optimal time lag = −0.7 s). We then asked participants to combine these two control strategies by playing the video game Pong^31^ (Fig. 3C), where the firing rate of an individual motoneuron controlled the vertical position of a paddle. Both participants were able to readily switch between control strategies to move the vertical paddle and hold it in place to score points (Video 5), achieving up to four consecutive hits.

Having identified multiple pairs of independent motoneurons, we next investigated if these could be used to move a computer cursor in two dimensions. Participant one could modulate the activity of two motoneurons, each mapped onto the movement of a cursor along a different direction, to achieve successful 2D navigation (Fig. 4A), and cursor control (Fig. 4B; Video 6). Targets requiring co-activation of both units were slightly more difficult to reach, as indexed by longer completion times (Fig. 4C); indeed, most of them were reached by activating the two motoneurons sequentially (Video 6).

Reanimating a participant’s paralysed hand would offer a more dramatic functional benefit than controlling a computer cursor given patients’ priorities^2^. We thus mapped participant one’s motoneuron activity to control a wearable soft exoskeleton that used motors and wires to flex his fingers upon motoneuron activation (Fig. 5A,B). By reassigning the motoneuron to different motors, he was able to actuate different fingers and achieve different grasp types. This allowed him, for example, to volitionally grasp a sponge (Fig. 5C) and a pen (Fig. 5D) by performing a tripod grip (Video 7).

**Fig. 5:**
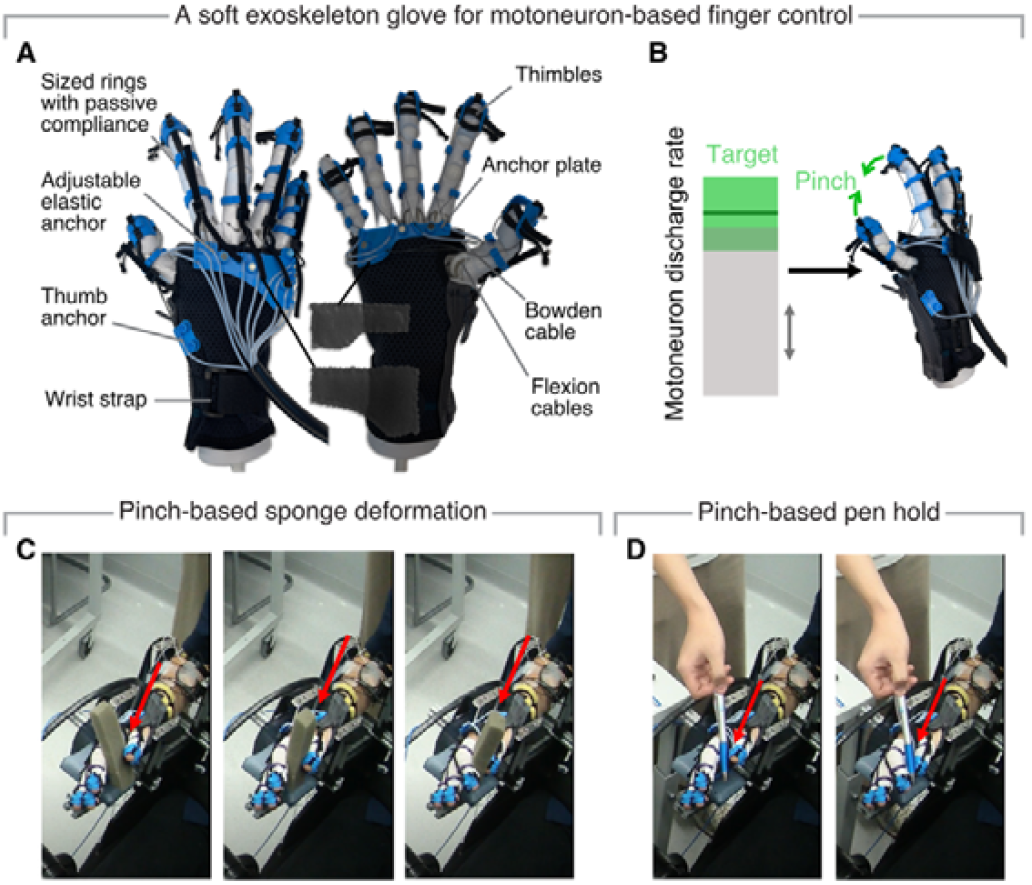
Reanimation through a soft exoskeleton. **A** The soft exoskeleton glove is operated via motors that pull on thimbles that are attached at the end of the fingers. **B** Firing of a selected motoneuron to a target level activates a combination of motors that pulls on the thimbles to achieve individual finger movements and grasps. The participant used motoneuron control to voluntarily deform a sponge (**C**) and grasp and hold a pen (**D**).

## Discussion

We have demonstrated one- and two-dimensional control of a computer cursor and a soft exoskeleton based on the activity of spinal motoneurons in two tetraplegic individuals detected from high-density intramuscular microelectrode arrays^24^. Our findings thus highlight the potential of intramuscular interfaces for restoring motor function in individuals with paralysis.

This work further provides insights for developing motoneuron-based interfaces. First, the value of recording from a large volume of muscle lies in being able to extract multiple control signals from a single implant, superseding conventional intramuscular^23^ and surface EMG approaches^5,32,33^. That said, we used microelectrode arrays whose recording length was 2 cm, which is likely smaller than the typical width of the extensor digitorum muscle^34^; future efforts could arrange electrodes over a longer length^18^ to optimally record from all its four bellies. Since each belly pulls on a different digit, this should afford four-dimensional control from a single implant.

Second, control was implemented with minimal processing of motoneuron activity: cursor displacement was proportional to the motoneuron discharge rate. This can be improved by adding a more complex mapping of motoneuron firing to control signals that accounts for their nonlinear response^35^, or by deploying adaptive algorithms inspired by work in intracortical brain-computer interfaces^36,37^. Interestingly, both participants reported difficulties with the *timing* of proportional control, citing a perceived mismatch between the onset of their attempt to move and initial movement of the cursor. Given that the cursor position was updated every 75 ms, which is below the usual ~100 ms threshold for EMG controller delay^38^, we think this mismatch arose from the non-linear relationship between an attempted motor command and the detected motoneuron action potentials^39^. Specifically, the movement of the cursor was possible only when the supraspinal input exceeded the motoneuron’s recruitment threshold; an effort below this threshold could have been perceived by the participant but did not cause motoneuron firing and cursor movement. Since combining the activity of multiple motoneurons is thought to provide a better estimate of the motor command input to a motoneuron pool^40^, multi-neuron mappings should help overcome this perceived delay.

Despite the promising results, there are limitations to this approach. Although minimally invasive, implantation carried risks such as infection and bleeding; these were mitigated by adhering to aseptic techniques, using ultrasound guidance and screening for blood-thinning medications. Furthermore, we foresee potential technological advancements that allow for full chronic implantation for years, going beyond the current limit of acute percutaneous implants that can remain in the muscle only for weeks^41–43^. From a control perspective, the number of independent control units is intuitively limited by the number of distinct (attempted) movements that the recorded muscles can generate. However, recent research indicates that combinatory movements, such as flexing two fingers together, may recruit distinct motoneurons^44^, which would increase substantially the number of available control signals.

In summary, our findings provide a proof-of-concept of an implantable system that uses high-density intramuscular microelectrode arrays to enable accurate, multidimensional control in individuals with tetraplegic SCI. Participants provided positive feedback on the intuitiveness of the control and expressed enthusiasm about using their own muscle signals, noting that this experience had a “beneficial effect on their mental health and overall well-being”. Overall, our results lay the groundwork for chronic motoneuron-based interfacing, with the potential to significantly improve the quality of life for individuals with paralysis arising from SCI, stroke and other neurological conditions.

## Methods

### Participants and ethical approval

Participant one was a male in his 60’s who suffered a spinal cord injury at C3, 14 months prior to participation in this study. He had no movement in his right upper limb (MRC grade 0/5) except for ring finger extension, which was rated MRC grade 1/5 (flicker of movement). The tone in the upper limbs was flaccid bilaterally and he had no function of his upper limbs.

Participant two was a male in his 60’s who suffered a spinal cord injury eight years prior to this study. His level of injury was at C2/C3. He had no function in his left hand. His left elbow and fingers were held in fixed flexion, and he had significant spasticity throughout his upper limb. He had no movement on attempted finger flexion or extension (MRC grade 0/5) and only flickers of movement on thumb extension (MRC grade 1/5). Ethical approval was provided by the NHS HRA (REC reference 23/SW/0140) and Imperial College Research Ethics Committees (ICREC reference 19IC5641), and was performed in accordance with the Declaration of Helsinki.

### Acquisition

#### A) Screening of candidate muscles using ultrasound

We targeted muscles with multiple tendinous compartments, reasoning that a single intramuscular microelectrode array in such muscles could capture activity from several attempted movements. This approach aimed to maximise the likelihood of detecting distinct motoneuron populations—and therefore independent control signals—from a single array. Given abnormalities in motor control after SCI^10^ (i.e., low-amplitude signals and abnormal synergies), ultrasound imaging (Butterfly iQ+) was used to identify areas of preserved muscle activity. During scanning, participants were instructed to attempt isolated flexion and extension movements of individual fingers, the wrist, and the thumb. Intramuscular electrode insertion was then targeted towards visible residual fibre contractions observed during these attempted movements.

#### B) High-density intramuscular electromyography

We recorded high-density intramuscular EMG using a 40-channel intramuscular electrode array^24^. Each electrode has an area of 5257 μm^2^ and are linearly distributed over a length of 2 cm. Microelectrode arrays were sterilised using ethylene oxide prior to insertion. The insertion site was first marked with a medical pen, after which the skin was sterilised used an alcohol wipe. An initial entry point was made with a 23G needle, to allow easier passage of the 25G needle threaded with the microelectrode array, comprising of the electrode contacts, and a guiding filament. Insertion of the microelectrode array was performed under ultrasound guidance into the muscle of interest. As an example, in the Extensor Digitorum Communis, the needle tip was guided laterally across the muscle belly (at an angle with respect to the horizontal of <30 degrees), from the compartment actuating little finger extension, to that of index finger extension. This in turn maximised the likelihood that the entire electrode contact span resided in the various muscle bellies attributed to individuated finger movement. Once the intended insertion path was confirmed, the needle was removed, leaving the microelectrode array in place. Insertion of the array was straightforward (taking around 30 minutes in laboratory conditions). High-density iEMG signals were then recorded in a monopolar montage using a multi-channel amplifier (Open Ephys FPGA Acquisition Board, 3^rd^ Generation) at 10 kHz.

#### C) High-density surface electromyography

We placed a 64-channel surface EMG grid (8 mm interelectrode distance) over the skin corresponding to the extensor digitorum muscle in both participants. In participant one, this was separately to the insertion of the microelectrode array whereas it was adjacent to the site of insertion of the microelectrode array in participant two. Signals were collected using the same Open Ephys amplified at 10 kHz. Ground and reference electrodes were also placed at the wrist.

### High-density EMG decomposition

We recorded high-density EMG during isolated extension tasks involving the fingers, thumb, and wrist, using both intramuscular and surface electrode arrays. For offline decomposition, participants were asked to follow a trapezoidal-shaped profile, with the plateau set to match their previously measured maximum voluntary contraction (MVC) for 20 seconds. During this task, they received real-time visual feedback based on the median root mean square (RMS) EMG signal across all channels.

Recorded signals were first band-pass filtered between 10–4400 Hz for intramuscular EMG, and between 10– 500 Hz for surface EMG. Following this, noisy channels were manually rejected, and the remaining data was extended and whitened. Extending and whitening the data permits better exploitation of the spatiotemporal dependencies in the EMG signal structure, removes second-order correlations, and normalises signal variance across channels^45^. We used Swarm-Contrastive Decomposition^26,46^ (SCD), a blind source separation method that recovers the activity of individual motoneurons by exploiting the spatiotemporal structure of motoneuron action potentials (MUAPs) and the sparsity of motoneuron firing. SCD employs an adaptive contrast function optimised through a particle swarm algorithm, allowing for enhanced separation of sources compared to methods using fixed nonlinearities. For this analysis, we assumed isometric conditions, stationarity of MUAP shapes, and a linear time-invariant EMG model.

To reduce computational load during experimental sessions, each high-density EMG file was initially decomposed using 10 iterations per array/grid. Motoneurons were retained if their silhouette (SIL) coefficient exceeded 0.85 (the SIL coefficient is a cluster validation metric used to quantify the separation between spike and noise centroids, as calculated by applying *K*-means clustering on extracted motoneuron pulse trains)^25^. After each iteration, motoneurons that fulfilled the SIL criterion were subtracted from the EMG signal. The EMG residual was then used as initialisation to the particle swarm optimiser for the following iteration, in a ‘peel-off’ strategy. The ‘peel-off’ approach facilitates the identification of additional motoneurons, particularly those with lower-amplitude or partially overlapping waveforms, by reducing temporal interference from previously extracted units. Following the experiment, all recordings were reprocessed offline using 50 decomposition iterations and with SIL set to 0.9.

Duplicate motoneurons were identified by pairwise comparison of spike trains using cross-correlation. Trains were temporally aligned within a ±50ms lag window, and similarity was quantified as the proportion of spikes co-occurring within a ±5ms jitter window. Units with a rate-of-agreement > 40% were classified as duplicates; the unit instance with the lowest coefficient of variation in inter-spike intervals within a given duplicate group was retained. Duplicate removal was performed both within and across arrays for high-density intramuscular EMG data but restricted to within-array/grid comparisons when intramuscular EMG and surface EMG data were concurrently recorded.

### Online control

To visualise real-time motoneuron firing, we applied motoneuron filters derived from offline decomposition to incoming EMG data^30^. However, because the offline process employed a peel-off strategy, the original filters—based on residual signals with reduced temporal overlap—were not directly applicable for an online paradigm. ‘Peel-off’ cannot be applied causally in an online setting without introducing substantial latency. Therefore, filters were recalculated directly from the raw EMG signal to reflect the non-idealised signal structure encountered during real-time operation. This recalibration reintroduces the temporal overlaps that were removed during offline decomposition, providing a more realistic basis for online spike detection. Below, we describe the filter recalibration procedure and its application to streaming EMG data.

#### A) Recalculation of motoneuron filters for online control

First, the original motoneuron filters were applied to the raw EMG data—using the same segment from the offline decomposition—to estimate motoneuron discharge times as they appear in the unprocessed signal. Spike-triggered averaging was then performed on the raw EMG to recalculate the motoneuron filters, capturing the full temporal overlap structure absent from the ‘peel-off’ residuals. These recalibrated filters were subsequently reapplied to the raw EMG to extract updated spike and noise centroids, along with normalisation factors that constrain the amplitude of the motoneuron pulse trains between 0 and 1. This final step is critical: filters derived from residual signals produce suboptimal projection magnitudes and noise distributions, leading to misaligned centroids and elevated false positive and false negative rates during threshold-based detection.

All recalibration steps were conducted using only the EMG channels retained during offline decomposition (i.e., excluding previously rejected channels). The resulting centroids and normalisation factors were used during online decoding to minimise computational demands, eliminating the need for real-time peel-off or k-means clustering on each incoming EMG batch.

#### B) Real-time online processing procedure

After the accommodations described above, online EMG processing followed previously established methods^30^. Incoming EMG batches were spatiotemporally extended proportional to the number of EMG channels, then whitened using the whitening matrix calculated on the previous offline data. Recalibrated motoneuron filters were applied to the whitened EMG, and the resulting pulse trains were normalised. Spike classification was performed by comparing each pulse train value to the corresponding recalibrated spike and noise centroids in Euclidean space; values closer to the spike centroid were classified as spikes. This approach eliminated the need for online k-means clustering, reducing latency in feedback generation. EMG data was processed in batches of approximately 75 ms. Although the batch duration varied slightly due to the acquisition system (Open Ephys, 3rd generation FPGA board), it did not exceed 125 ms.

Firing rates were computed as the number of detected spikes within a sliding window—800-1000ms for both 1D and 2D tasks—divided by the window length. These bin sizes were chosen to reflect task design requirements and to ensure appropriate temporal smoothing.

Tracking performance for the 1D dynamic tasks was calculated by first determining the optimal temporal alignment (i.e., time lag) between the target and the discharge timings of the motoneurons. We systematically tested time lags from –1.5 s to 1.5 s in 200 ms increments to find the alignment that optimised a combined performance metric (70% correlation and 30% NRMSE). The metrics were then computed using the temporally aligned signals.

#### C) Combining motoneuron filters and whitening across movement types

Since participants performed each intended movement separately, the corresponding offline EMG recordings produced distinct feature sets, including different whitening matrices for each movement. However, during online control, only a single whitening matrix can be applied to the incoming EMG data, regardless of the movement being performed. To ensure consistent preprocessing with an online decomposition that whitens appropriately across all movements, a unified whitening matrix was computed.

This was achieved by concatenating preprocessed EMG data segments from all recorded movements and then applying whitening to the combined dataset. This approach ensured that the resulting whitening matrix accounted for the statistical variability across all movement types. Finally, detailed above, the centroids and normalisation parameters were subsequently recalibrated to ensure compatibility with the unified whitening matrix during online application.

### Online control paradigms

#### A) Real-time raster

We sought to assess whether individual motoneurons could be controlled online. To select an appropriate motoneuron for control, we displayed the online firing rate of motoneurons that were identified during decomposition, whilst participants attempted individual finger, thumb and wrist movements (i.e., we showed participants the online firing rate of ring extension motoneurons while they attempted ring extension). The motoneuron to be controlled was chosen by the study investigators and the participant, based on an impression of which one was most under voluntary control; that is, a motoneuron that fired upon attempted movement but did not fire during rest, and which increased its firing rate proportionally with increasing effort.

#### B) One-dimensional profile tracking

A virtual cursor was controlled by participants, whose position change as a function of the firing of a single motoneuron, as they tracked the following profiles:

1. Trapezoid – increasing firing rate to 50% of the maximal firing rate during decomposition and sustaining it for 20 seconds.
2. Dynamic – made from four distinct sinusoidal segments, each covering two complete cycles. These segments vary in frequency, either 0.25 Hz or 0.5 Hz, and in amplitude, corresponding to either 25% or 50% of the maximal voluntary contraction (MVC). Importantly, firing rate returns to 0 between cycles.

#### C) Pong

In this task, participants vertically controlled a paddle to intercept a ball bouncing within screen boundaries. The task was designed to combine elements of one-dimensional tracking of a trapezoidal profile—primarily assessing sustained output of motoneuron firing—with aspects of dynamic tracking—the ability to modulate the temporal patterning of motoneuron firing. Paddle position was driven by the real-time firing rate of a selected motoneuron, with higher rates producing greater upward movement. Participants scored one point per successful interception and lost one point for each miss. The score was continuously displayed to maintain engagement.

Task difficulty was modulated by adjusting ball speed, paddle size, and the maximum firing rate threshold, which set the upper bound of paddle position; lowering this threshold reduced the required muscle activation to reach the top of the screen. Thresholds were individually tuned to balance challenge and controllability across participants. Ball trajectories were semi-randomised: each reset positioned the ball at the centre with a trajectory directed away from the paddle, allowing time for movement preparation.

#### D) Two-dimensional control

We used a two-dimensional (2D) quadrant interface to evaluate the feasibility of two-dimensional motoneuron control. Each axis of the quadrant was parameterised by the firing rate of a motoneuron that could be volitionally and independently modulated. These motoneurons were selected based on the online raster, and their filters were recalibrated to account for both ‘peel-off’ effects and the concatenation of motoneurons across all movements identified offline.

Targets were presented either along a single axis, requiring isolated activation of one motoneuron (x-axis for the first unit, y-axis for the second), or off-axis, necessitating concurrent activation of both units in varying combinations of firing rates. Target locations were generated pseudorandomly within an area bounded by the origin (both motoneurons not firing) and the maximum of each motoneuron’s firing rate. Targets had a tolerance window of ±30% whilst the firing rate binning window was 800 ms, allowing for a more stable estimate of firing rate and better capturing volitional variability, beyond that of single-spike contributions.

To discourage transient or incidental passage through the target region and promote stable, intentional control, a hold requirement was imposed. The firing rates of both motoneurons had to remain within the target tolerance window for a minimum duration of five EMG update intervals (375 ms) for the trial to be considered successful. Upon successful completion, participants were instructed to return to a resting state before the next target was presented.

### Soft exoskeleton control

A soft exoskeleton glove was used to demonstrate physical control in participant one. This was not possible in participant two due to significant spasticity resulting in fixed flexion of the fingers. The cable-driven exo-glove enables active finger flexion via motors, and passive extension via springs. The glove offers up to five independent degrees-of-freedom, which can be combined to perform several functional movements, allowing users to interact with a variety of objects (demonstrated using a pen, syringe, foam block, and a thin wallet).

The glove is built upon a soft zip-on/off brace, which serves as a base and allows easy donning/doffing. A semi-flexible 3D-printed (PLA) plate is attached to this base via snap-fasteners, serving as an anchor and guide for the Bowden housing that route the cables to the actuation pack. Rings and thimbles transmit power from the actuation pack to the fingers through cables for flexion and straps for extension, respectively.

The glove was designed for easy donning and doffing, offering a finger-first slip-on design. Critically, the glove was designed to be highly modular and customisable to accommodate subject variability – with all components able to be swapped on-the-fly. Rings and thimbles were sized for the patient (from a selection of diameters 12-20 mm), with mechanical compliance to account for short-term fluctuations during experiments, *e*.*g*. from swelling. Additional finger padding (white bandages) was applied beneath the glove as a precautionary measure.

Cables from the glove were connected to the motors via a second set of weaker cables, which acted as a mechanical fuse and limited force transmission to the fingers. Dynamixel MX-128 and MX-64 motors (Robotis Co. Ltd., South Korea) were daisy-chained together and connected to the PC running the control algorithm. Current-based position control mode was used to limit applied torque while allowing control over joint velocities and joint positions. For compound movements, joint velocities were automatically set to achieve pinch, tripod and grasp movements.

The detected motoneurons were assigned to different finger or compound movements during the experiments. The extension to fully-open position post-flexion movements were made passively.

## Data availability

Videos can be found on Zenodo (https://doi.org/10.5281/zenodo.15870327). Data will be made available upon reasonable request to the corresponding author.

## Code availability

The code for analysis is available from the corresponding author upon reasonable request.

## Author contributions

A.G., C.G., V.R. and L.G.V. collected and analysed data. A.G., C.G., V.R., J.A.G. and D.F. wrote the paper. I.S., R.V. and P.B. supplied the soft exoskeleton and provided support with its use. A.K. provided hardware support. X.Y. provided technical support. A.G., C.G., V.R., J.A.G. and D.F. conceptualised the study. J.A.G. and D.F. supervised the study.

## Supplementary Figures

**Suppl. Fig: 1.**
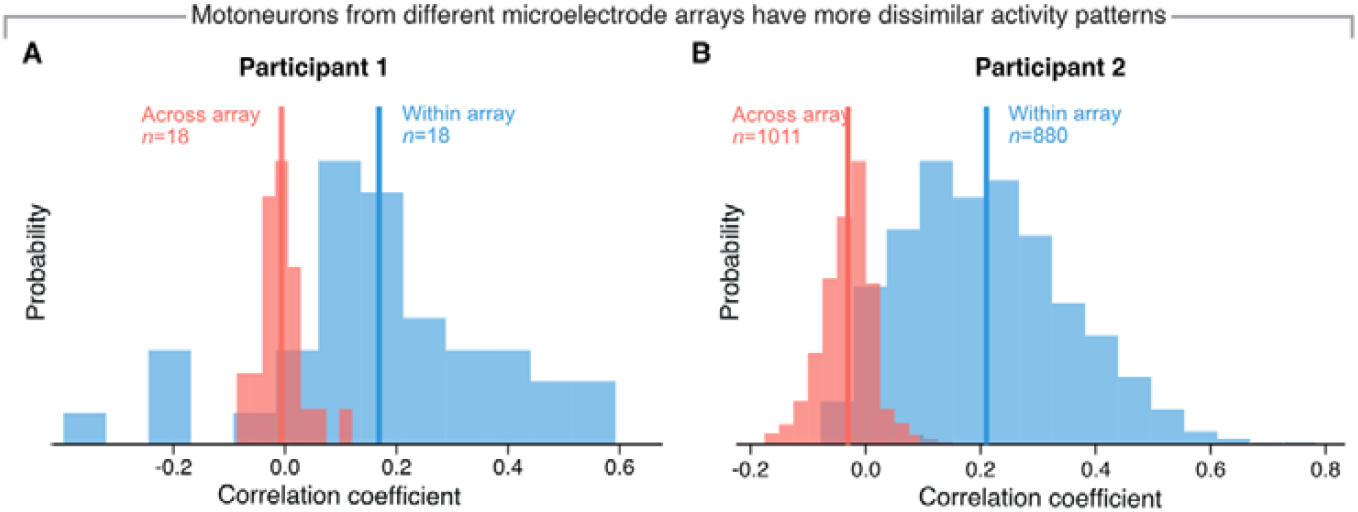
Motoneurons from the same microelectrode array tend to have more correlated activity than motoneurons from different microelectrode arrays. Correlation coefficients are computed between motoneuron firing rates within a single microelectrode array or across microelectrode arrays for participant one (**A**) and participant two (**B**).

**Suppl. Fig: 2:**
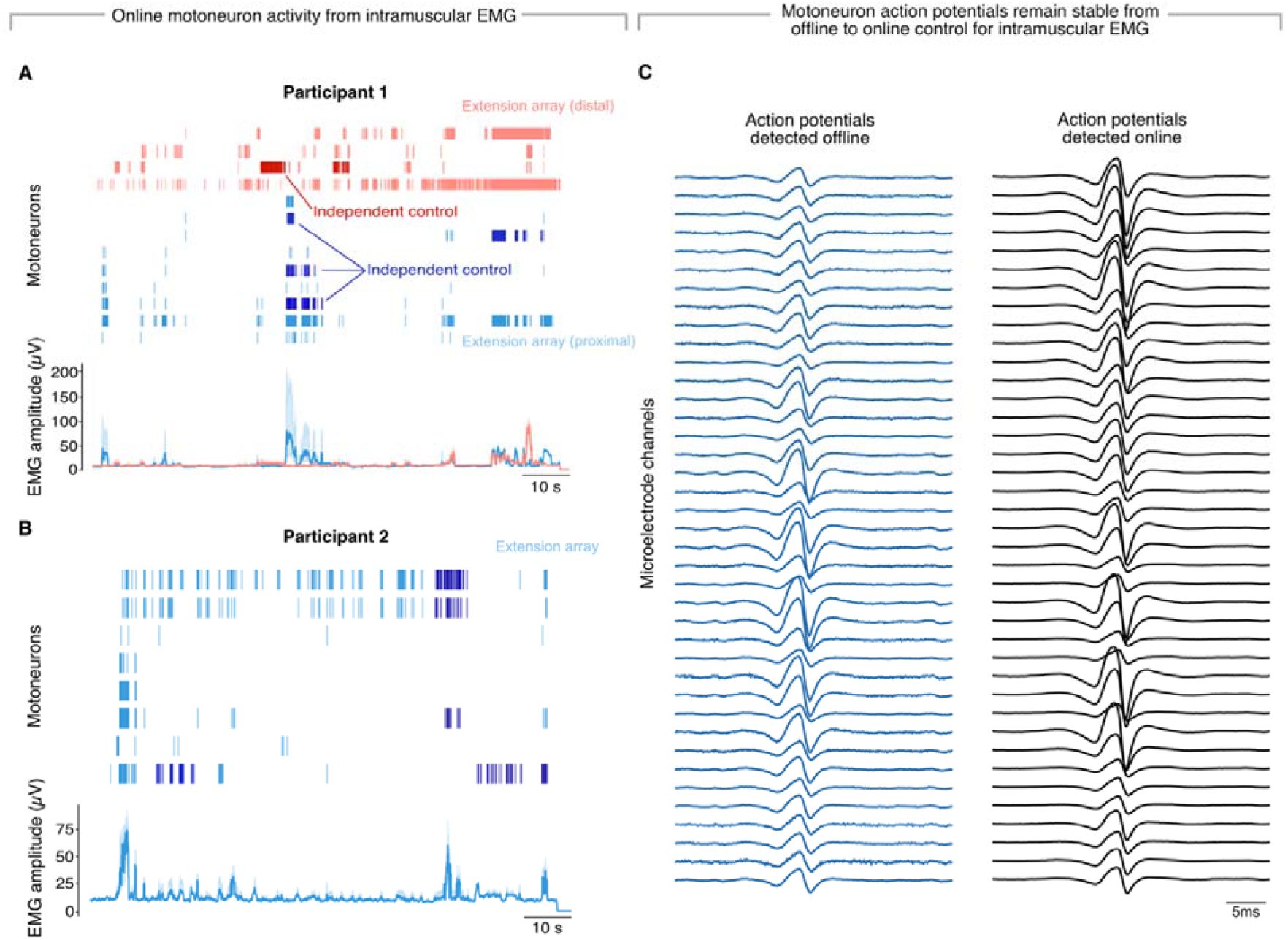
Online raster plot of identified motoneurons. Raster plots show real-time activity of individual motoneurons during different attempted movements from participant one (**A**) and two (**B**). Bottom plots show overall EMG amplitude from each array, calculated as the root-mean-squared value. Examples of motoneurons that fired independently of one another during different attempted movements are highlighted. A subsample of all motoneurons were displayed back to each participant to facilitate ease of feedback. Stability of a motoneuron action potential shape between the offline and the online experimental session (**C**) over a four-hour period. The displayed motoneuron was one of the two independent ones used in the 2D control by participant one.

**Suppl. Fig. 3:**
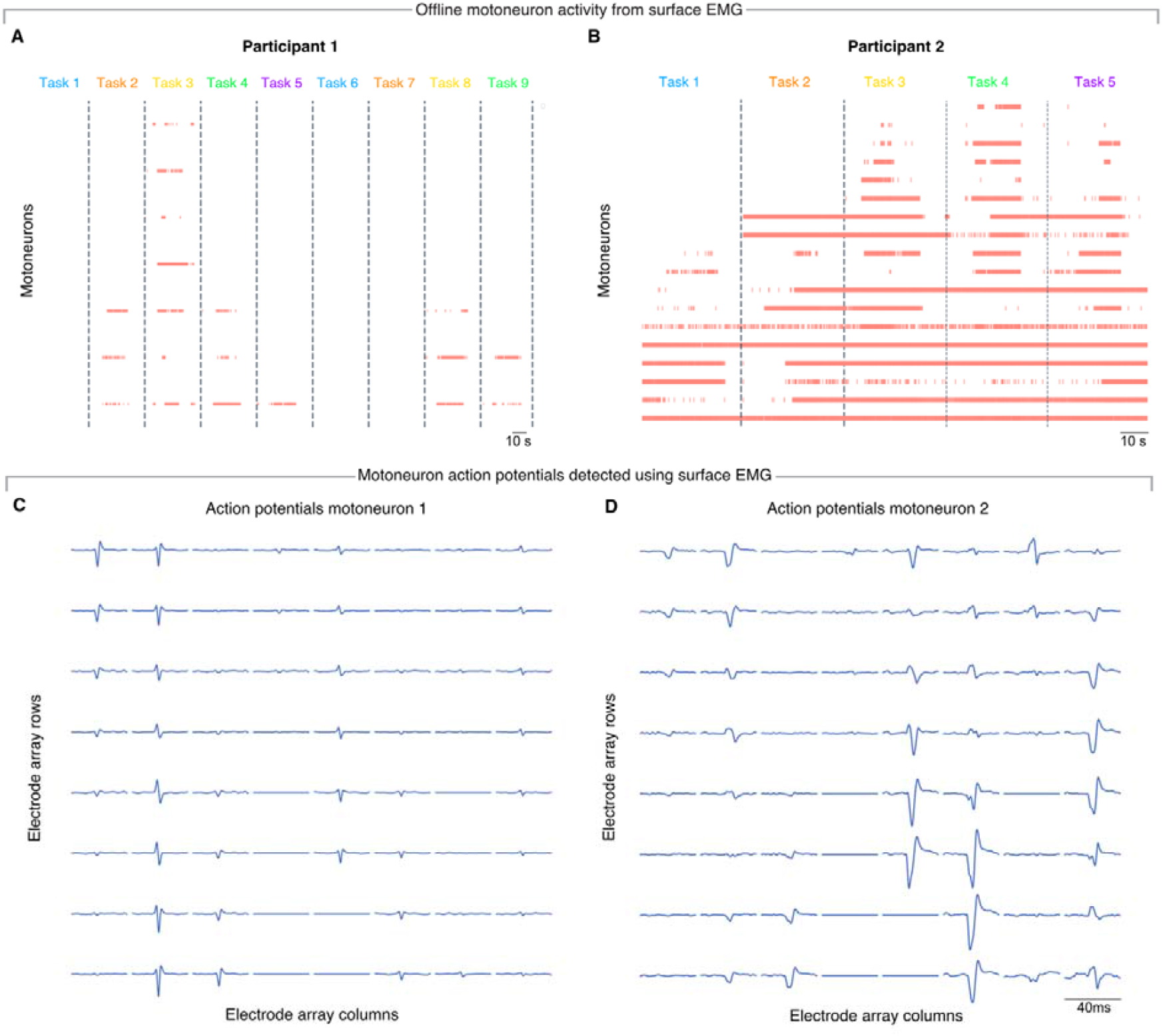
Motoneurons identified using high-density surface EMG. Motoneurons identified offline during different attempted movements for participant one (**A**) and two (**B**). No independent motoneurons were identified for participant one, two different pairs of motoneurons were identified as independent for participant two. Examples of motoneuron action potential shapes across the 64-channel surface EMG grid detected in participant two (**C**).

**Suppl. Fig. 4.**
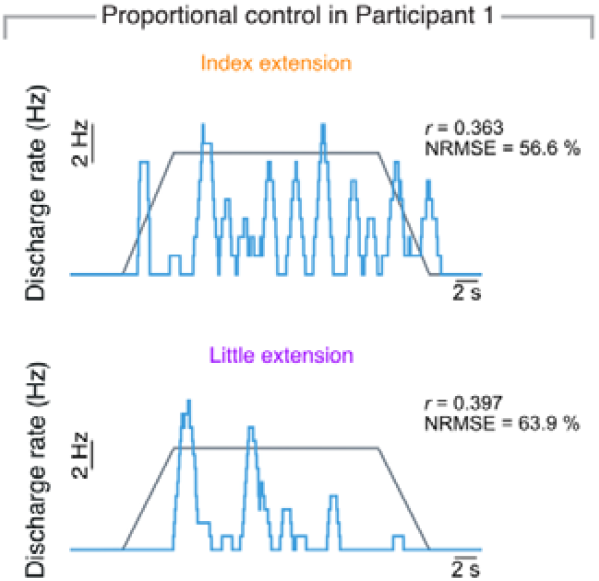
Additional one-dimensional tracking data. Online tracking of a trapezoidal profile using the firing rate of two different motoneurons each associated to a different attempted action by participant one.

